# Gamified Adaptive Approach Bias Modification in Individuals with Methamphetamine Use History from Communities in Sichuan: A Pilot RCT

**DOI:** 10.1101/2023.08.22.22279466

**Authors:** Danlin Shen, Jianping Jiao, Liqun Zhang, Yanru Liu, Xiang Liu, Yuanhui Li, Tianjiao Zhang, Dai Li, Wei Hao

## Abstract

**BACKGROUND:** Cognitive bias modification (CBM) programs have shown promise in treating psychiatric conditions, but they can be perceived as boring and repetitive. Incorporating gamified designs and adaptive algorithms in CBM training may address this issue and enhance engagement and effectiveness.

**OBJECTIVE:** To gather preliminary data and assess the preliminary efficacy of an adaptive approach bias modification (A-ApBM) paradigm in reducing cue-induced craving in individuals with methamphetamine use history.

**METHODS:** A randomized controlled trial with three arms was conducted. Individuals aged 18-60 with methamphetamine dependence and at least one year of methamphetamine use were recruited from 12 community-based rehabilitation centers in Sichuan, China. Individuals with inability to fluently operate a smartphone and/or the presence of mental health conditions other than methamphetamine use disorder (MUD) were excluded. A-ApBM group engaged in ApBM training using a smartphone application for four weeks. A-ApBM used an adaptive algorithm to dynamically adjust the difficulty level based on individual performance. Cue-induced craving scores and relapse were assessed using a visual analog scale at baseline, post-intervention, and at week-16 follow-up.

**RESULTS:** A total of 136 participants were recruited and randomized: 48 were randomized to the A-ApBM group, 48 were randomized to the S-ApBM group, and 40 were randomized to the no-intervention control group. The A-ApBM group showed a significant reduction in cue-induced craving scores at post-intervention compared to baseline (Cohen’s d = 0.34, p < 0.01, 95% CI = [0.03,0.54]). The reduction remained significant at the week-16 follow-up (Cohen’s d = 0.40, p= 0.01, 95% CI = [0.18,0.57]). No significant changes were observed in the S-ApBM and control groups.

**CONCLUSION:** The adaptive ApBM paradigm with gamified designs and dynamic difficulty adjustments may be an effective intervention for reducing cue-induced craving in individuals with methamphetamine use history. This approach improves engagement and personalization, potentially enhancing the effectiveness of CBM programs. Further research is needed to validate these findings and explore the application of adaptive ApBM in other psychiatric conditions.

**TRIAL REGISTRATION:** Registered at clinicaltrials.gov (ID NCT05794438).

## Introduction

Cognitive biases, such as attention bias, approach bias, and interpretation bias, are intricately linked to the psychopathologies of conditions, including anxiety, depression, post-traumatic disorders, and addiction. Typically, individuals with psychiatric disorders exhibit a distinctive cognitive bias towards certain types of stimuli. Researchers have developed cognitive bias modification (CBM) training schemes aimed at redirecting the bias from negative to positive stimuli. The mechanisms underlying CBM’s efficacy are grounded in theories of automaticity and habit formation, where repeated exposure to corrective training diminishes the influence of negative stimuli over time. For example, pertaining to addiction, approach bias modification trainings aim to correct the cognitive bias of approaching addictive substances; and for treating anxiety and depression, attention bias modification and interpretation bias modification aim to correct the cognitive bias of paying excessive attention to negative emotions and interpretations. CBM interventions have shown potential in treating addiction, anxiety, and depression. [1–2]

CBM training, although effective, has been criticized for being monotonous and unengaging.[3] Several studies have reported the issue of boredom with CBM training.[4–6] Qualitative interviews with patients [5] have revealed that participants often found CBM to be "boring," "repetitive," "tedious," and that they "just tried to get through it as quickly as possible." This may be attributed to the nature of CBM’s design, where each session typically consists of repetitive trials, and the rules and instructions remain the same throughout the intervention program. Consequently, the tedium and boredom associated with CBM training may result in a loss of attention and interest in the program, thereby hindering its efficacy. However, given the evidence of the effectiveness of CBM in treating various psychopathologies, it is crucial to address the issue of boredom and make the training more engaging and fun for participants to further enhance efficacy.

In treating substance user disorders, Approach Bias Modification (ApBM) is a type of CBM training method aimed at counteracting approach biases and ultimately reducing the craving and usage of addictive substances. Through repeated exercises, participants are exposed to substance-related images and trained to make avoidance movements (e.g., pushing an image away) while associating non-substance-related images with approach movements (e.g., pulling the image closer). This retraining helps shift automatic tendencies, encouraging avoidance of substance-related cues. Research, including several randomized controlled trials [7–11], has shown that incorporating ApBM into treatment programs for substance use disorders can reduce relapse rates, even with brief training sessions of 10–15 minutes conducted over multiple sessions.[11]

To make CBM training more engaging and effective, efforts have been made to incorporate gamification into CBM, with some studies demonstrating improved engagement and outcomes through the use of animations, sounds, feedback, and point-scoring systems [12]. The integration of gamification in digital therapeutics, particularly in CBM, is becoming more prevalent, suggesting a promising avenue for enhancing the efficacy of these interventions [12–15]. This study introduces an innovative approach: an Adaptive Approach Bias Modification (A- ApBM) scheme designed to counter the monotony of traditional CBM. This scheme not only includes gamified elements but also features an adaptive algorithm that adjusts the training difficulty based on individual performance to enhance engagement.[16,17] The key idea of Adaptive Approach Bias Modification (A-ApBM) is to use an algorithm to dynamically estimate individual performance and adjust the training difficulty accordingly. In contrast to traditional Static Approach Bias Modification (S-ApBM), where the difficulty level is fixed throughout the intervention, A-ApBM continuously adapts to maintain an optimal level of challenge. This ensures personalized engagement, keeping participants motivated and maximizing the intervention’s effectiveness. Difficulty adjustment is a crucial concept in game design because it provides participants with a sense of challenge that matches their skill level. By striking a balance between ease and difficulty, the intervention avoids boredom from being too predictable and frustration from being too demanding. This sense of challenge promotes a state of "flow," where participants are fully immersed and engaged in the task. By making the training feel more like a dynamic and rewarding game, A-ApBM not only enhances the overall experience but also increases adherence and improves therapeutic outcomes. This ensures personalized engagement, keeping participants motivated and maximizing the intervention’s effectiveness. This is expected to maintain engagement and challenge throughout the program. While previous studies have attempted to modify CBM difficulty for better outcome, our approach is distinguished by its analytical and quantitative emphasis on algorithm-based adjustment.[18.19]

The study is designed to be a randomized trial with three arms: the A-ApBM as the intervention arm, a Static Approach Bias Modification (S-ApBM) as the active control arm, and a no-intervention control arm. The inclusion of the S-ApBM group was crucial for evaluating the effectiveness of the adaptive intervention. The S-ApBM group served as an active control to isolate the impact of the adaptivity—the dynamic difficulty adjustments in the A-ApBM group. Both groups participated in bias modification training, but the S-ApBM group’s static nature allowed us to assess whether adaptivity specifically contributed to greater reductions in cue-induced cravings. This control also accounted for general engagement effects, ensuring that any observed differences were due to the adaptive algorithm rather than task participation alone. By including both versions, we tested whether personalized, adaptive training improved outcomes compared to static training, thus helping to clarify the role of adaptivity in enhancing cognitive interventions. Given the significant public health burden posed by methamphetamine addiction [20–23], this study’s findings may inform the development of more effective therapeutic tools that engage users and sustain their motivation throughout treatment.

## Method

### Study Design

This study was a three-armed randomized controlled trial, including one intervention group (A-ApBM) and two control groups (a S-ApBM group as active control and a no-intervention control group). Participants were randomly assigned (using computer-generated random numbers) to the three groups using a computer random number generator. This method ensured that each participant had an equal chance of being placed in any of the groups. While the trial incorporated blinding, due to the presence of a no-intervention control group, it is more accurately described as partially double-blinded. Specifically, the participants in the intervention groups were unaware of whether they were receiving adaptive or static training, which helped reduce bias from the participant side. However, those in the no-intervention control group likely knew they were not receiving an active treatment, which limits full blinding. The participants in the A-ApBM and S-ApBM groups were asked to engage in daily trainings for four weeks.

### Ethical Considerations

This study was conducted in accordance with the Declaration of Helsinki and approved by the Ethics Committee of the Second Xiangya Hospital of Central South University (2021- 076) and is registered at clinicaltrials.gov (NCT05794438). Informed consent was obtained from all participants prior to their inclusion in the study. Participants were provided with detailed information about the study’s purpose, procedures, potential risks, and benefits, and they were informed of their right to withdraw at any time without consequence. All data collected during the study were anonymized before analysis. Participants’ identifiable information was securely stored and accessible only to authorized personnel. De-identified datasets were used for all analyses to ensure the protection of participants’ privacy.

### Sample Size Calculation

The sample size for this pilot trial was determined based on practical considerations rather than formal statistical power calculations, as there was insufficient data to estimate the effect size and variance of the proposed A-ApBM intervention. We aimed to recruit 150 participants (50 per group), based on preliminary communications in December 2022 with 12 community rehabilitation centers. These centers’ rosters indicated that approximately 150 individuals met the inclusion criteria for the study. This sample size allowed for preliminary analyses and to assess the feasibility of the study procedures and intervention, while acknowledging the logistical constraints associated with recruitment, time, and resources.

### Participants

Participants were recruited from 12 community-based rehabilitation centers in Sichuan, China, all having a history of methamphetamine use and undergoing community-based rehabilitation. Participants were individuals aged between 18 and 55 years with a history of methamphetamine use for at least one year. The diagnosis of methamphetamine use disorder (MUD) was confirmed based on community rehabilitation center records and self-reports.

Additionally, we excluded individuals with any other mental health disorders, as well as those who could not fluently operate a smartphone, ensuring all participants could engage with the smartphone-based intervention. These revisions provide a clearer understanding of the participant demographics and eligibility criteria. Participants’ characteristics were collected using survey, which was available in online Appendix 1.

### Intervention: A-ApBM

The A-ApBM group underwent a specialized intervention using the Wonderlab Harbour smartphone application (Adai Technology (Beijing) Co. Ltd.) for four weeks. This intervention is rooted in the principles of approach bias modification training, as delineated in previous studies on substance use disorders [10,11]. In each session, participants were instructed to swipe upward (or downward) when shown images in portrait (or landscape) orientation. After swiping upward (downward), an animation either shrank (grew) to create the effect of distancing (approaching) the object. The images depicted either methamphetamine-related items (such as crystals, powders, or paraphernalia) or representations of healthy living (e.g., wealth, sports, gourmet food, family activities, etc.).

The A-ApBM group’s training focused on congruency, as per Kruijt et al.’s model [24], where congruent trials aligned with the training’s intention, such as avoiding drug cues. Our study adopts a modified approach for calculating the Intended Training Ratio (ITR), using the ratio of congruent trials to the total number of trials. This adjustment was made to enhance mathematical robustness and to directly correlate the ITR with the training’s intensity and specificity. In A-ApBM, the ITR dynamically varied based on an algorithm (described in more detail later) that monitored the user’s performance, adjusting the difficulty index and ITR accordingly. This adaptive method is intended to individualize the training’s intensity to the user’s performance level, offering a personalized and dynamic approach to cognitive bias modification.

### Controls: S-ApBM Active Control and No-intervention Control

The study included two control groups: the S-ApBM group and a no-intervention control group. Participants in the S-ApBM group also used the same app for a similar duration but experienced a static form of ApBM training. Each session in the S-ApBM group consisted of a constant number of drug and non-drug trials, with a high proportion of congruent trials, resulting in a stable ITR of 92.3% (144 out of 156 trials) throughout the program. This static training model provided a control baseline to compare against the dynamic, adaptive A-ApBM intervention. The no-intervention control group did not receive any interventions, serving as an additional baseline to measure the effects of engaging in any structured intervention versus no intervention.

## Design of the A-ApBM Algorithm

### Algorithmic Components and Metrics

Intended Training Ratio (ITR): The ITR is crucial in the A-ApBM, representing the proportion of trials within a session that align with core training objectives, specifically avoiding drug cues and approaching non-drug-related cues.

Calculated as the ratio of congruent trials to total trials, this measure directly correlates with training intensity and contingency.

Performance Index: This index reflects the participant’s ability to meet the training’s objectives. Defined as 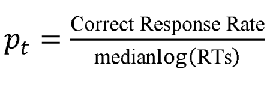 median of the logarithmically transformed response times, offering a comprehensive measure of a user’s performance in an ApBM training session t (the index *t* indicates the sequential session number within the intervention. The first training session would have t = 1, and the second training session would have t = 2, and so on).

Difficulty Index: We hypothesize that the perceived difficulty of an ApBM session by users is influenced mainly by two factors: the ITR and the variation in the ITR across sessions. The relationship between the ITR and difficulty is proposed to be an inverted U-shape. This shape is chosen because it effectively captures the nuanced way in which the ITR influences the cognitive processing required during the task. At an ITR of 100% or 0%, the task becomes predictable, with all trials being congruent, allowing users to form automatic associations between the stimulus type and format. However, at an ITR of 50%, where half of the trials are congruent and the other half are not, the task reaches peak difficulty. The 50% ITR creates maximum randomness in stimulus-type and format associations, making it challenging for users to discern a clear pattern, thus requiring more cognitive effort to perform correctly. The second crucial factor is the variability of the ITR in consecutive sessions. This aspect of difficulty is grounded in the concept of cognitive adaptability. If the ITR remains constant (or shows minimal variation) across sessions, participants may quickly adapt to the pattern, reducing cognitive load and perceived difficulty. In contrast, a high variation in the ITR across sessions introduces unpredictability, challenging the participant to continuously adapt to new patterns. This unpredictability requires greater cognitive flexibility and is hypothesized to increase perceived difficulty. To incorporate this factor, our model includes the standard deviation of the ITRs from recent sessions (SD(ITR_t-k+1_ ,… , ITR_t_)). A higher standard deviation indicates greater variability in ITR from one session to the next, thus contributing to an increased difficulty index. This dynamic adjustment ensures that the training remains challenging yet achievable, adapting to the individual’s learning curve and maintaining engagement throughout the intervention. To model the difficulty level, we utilize a quadratic term (ITR squared): 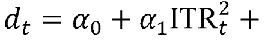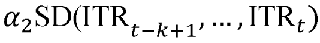, where a’s are to be estimated dynamically.

### Relationship Between Performance and Difficulty

The adaptive algorithm within A- ApBM is designed to learn a dynamic relationship between the participant’s performance index and the difficulty index. We employ a linear model to model the relationship between performance p_t_ and the difficulty index d_t_ in session t. Specifically, for each user i, we model the relationship using the equation 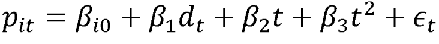. The terms 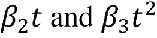 account for the nonlinear (quadratic) learning curve as the user furthers into the intervention program and becomes more familiar with the ApBM training. The term E_t_ represents the idiosyncratic error or residual error, which captures the random shocks that can affect performance.

### Sketch of the Algorithm

The A-ApBM algorithm initiates with three warm-up sessions, during which no ITR adjustments are made. Performance and difficulty indices for each participant are dynamically calculated at the end of each session, starting from the fourth session. From this point, a linear regression model is fitted to each user’s cumulative data, estimating the relevant parameters (as and (Js). Based on these estimations, the algorithm then adjusts the difficulty level of subsequent sessions to align with a predetermined difficulty curve. Building upon the analysis presented by Li et al. [12], four difficulty curve shapes are considered: U-shape, inverted U-shape, N-shape, and inverted N-shape. For our study, we have identified an inverted U-shaped difficulty curve as the most optimal for long-duration games with a general reward structure. This curve provides a balanced challenge, gradually increasing in difficulty to a peak before tapering off, effectively maintaining user engagement and motivation. A proprietary algorithm is employed to ensure that the user’s difficulty level closely follows this inverted U-shaped trajectory, adapting the training intensity in real time to the user’s performance and learning curve. An implementation of the A-ApBM algorithm is available at https://github.com/liuxiang91/A-ApBM/.

## Outcome Measures

### Primary Outcome: Self-Reported Cue-induced Craving Score

Self-reported cue-induced craving scores were utilized as the primary outcome measure. Participants were asked to review images that depict methamphetamine crystals, powders, and paraphernalia, and rate their cravings using a 0-100 visual analog scale. To improve the accuracy and reliability of participants’ self-reported cravings, they were instructed to rate the images twice upon enrollment: first on a smartphone and then again, one day later, on a printed paper. The average score of these two assessments was used as the baseline measure. The follow-up assessment took place in week 16 (12 weeks after intervention completion).

### Secondary Outcome: Relapse

Relapse was defined as the resumption of methamphetamine use following a period of abstinence. This was monitored through self-report and urine drug screening tests at week 4 and week 16 of the study. Participants were asked to provide urine samples at regular intervals during the follow-up period, which were tested for methamphetamine metabolites. Additionally, they were required to report any drug use incidents, including the date, amount, and context of use.

## Statistical Analysis

Data were analyzed using a mixed-design Analysis of Variance (ANOVA), with time as the within-subjects factor and group as the between-subjects factor. The dependent variable was the mean craving score. The ANOVA adhered to an Intention-to-Treat (ITT) approach, with missing outcomes imputed using the Last Observation Carried Forward (LOCF) method. To address the skewed distribution of craving scores and ensure robustness, a nonparametric bootstrap with 10,000 resamples was applied to calculate F statistics and effect sizes for pairwise comparisons. Confidence intervals were derived using the bias-corrected and accelerated (BCa) method.

For relapse, we also used an ITT approach by assuming the missing observations meant repalse. A 3-sample test for equality of proportions (using the prop.test function in R 4.2.1) was conducted by assuming the missings to have relapsed. The p-value was 0.23, indicating no significant differences.

To assess the proposed relationship between performance and difficulty indices, their association was retrospectively analyzed using data from the A-ApBM group. The performance index for the ApBM sessions was calculated. For the computation of the standard deviation of the past three ITRs, a rolling window of three was utilized, excluding the initial three sessions due to their constant ITRs. A linear mixed model was fitted to predict the performance index, incorporating ITR, ITR squared, *t* and *t* squared, and Past3ITRSD as predictors. The model accounted for subject variability through a random effects parameter.

All statistical analyses were performed using R 4.3.2. Significance levels for all tests were set at an alpha of 0.05.

## Results

### Participants and Participants Flow

In March 2023, 136 participants were screened and assessed for eligibility, and all participants were eligible and randomized into three groups using a computer random number generator: the A-ApBM group (n = 48), the S-ApBM group (n = 48), and the no-intervention control group (n = 40). The trial aimed to recruit 150 participants, however, after successfully recruiting 136 participants, the pool of eligible individuals from the centers’ rosters was exhausted. No new participants were available within the short-term timeframe, making further recruitment infeasible. Given that we had nearly reached our target and that 50 participants per group was considered sufficient for feasibility and preliminary analysis, the recruitment phase was concluded at 136 participants. Throughout the course of the study, dropout rates were monitored. All 136 participants completed the intervention (or control) at week 4. At week 16, three A-ApBM participants dropped out and were not evaluable due to missing follow-up outcomes. The no-intervention control group had two dropouts whose outcome were not evaluable at the 16-week follow-up. The S-ApBM group maintained full participation throughout the intervention phase. Overall, the dropout rate was 4% (5 out of 136). Table 1 presents the baseline characteristics of the participants by group. We conducted statistical tests among the groups. Specifically, Pearson’s chi-square tests were applied for categorical variables (e.g., sex, dominant hand, marital status, education, smoker, drinker), and the Kruskal-Wallis rank sum test was chosen for continuous variables (e.g., age, meth use history, craving at baseline) due to the non-normality of the data. As indicated in the table, all p-values are above 0.05, demonstrating that there are no statistically significant differences in participant characteristics across the three groups.

**Table 1:**
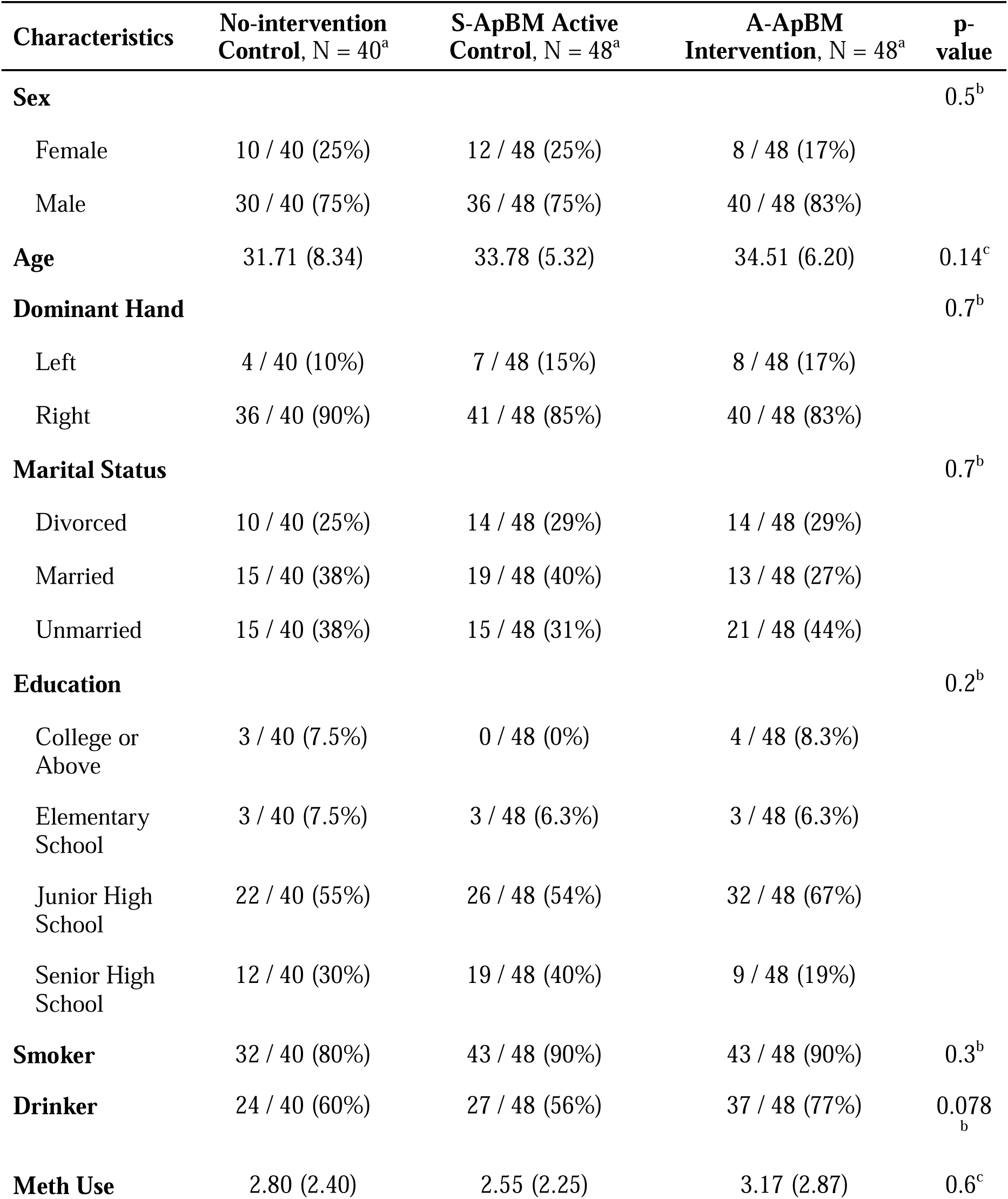
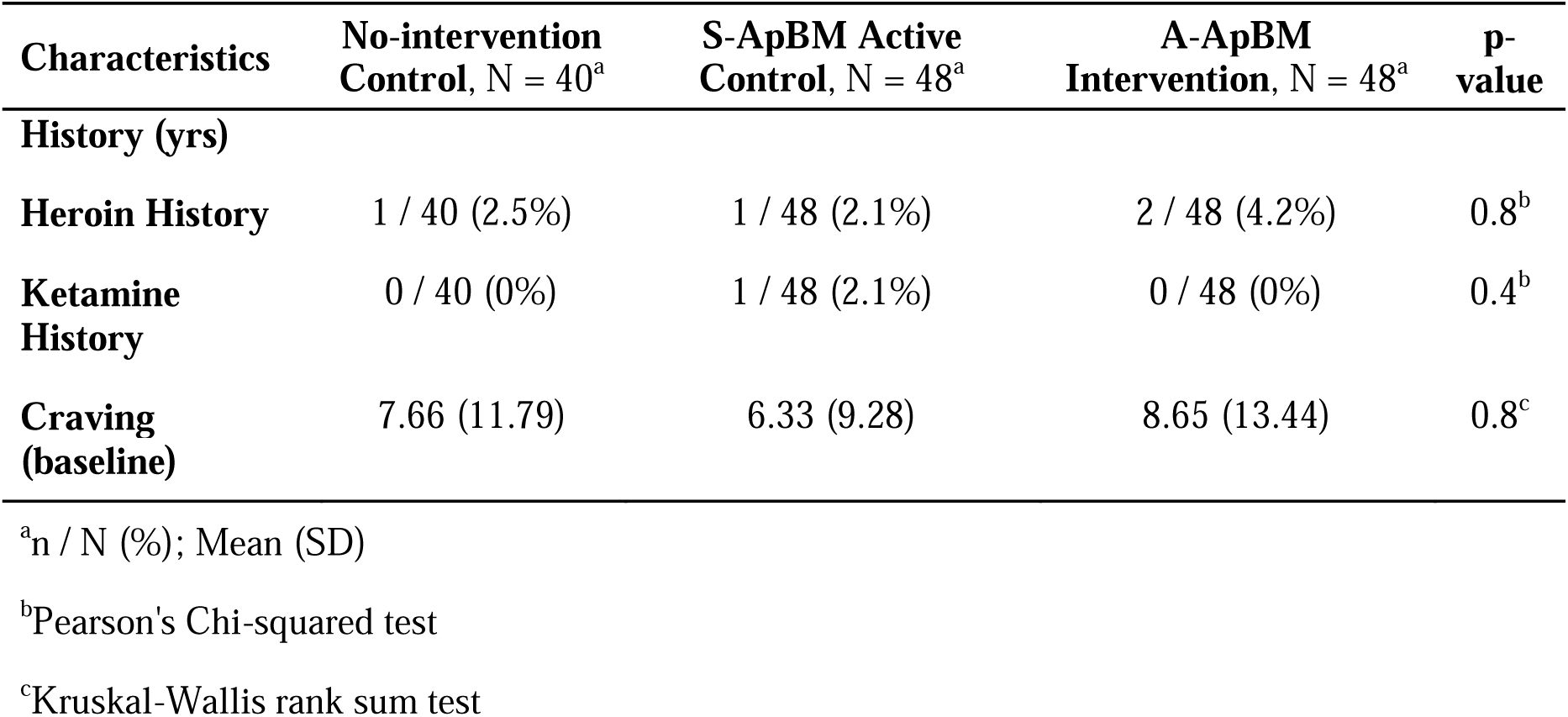
Participant Baseline Characteristics.

| **Characteristics** | **No-intervention**<br>**Control, N = 40<sup>a</sup>** | **S-ApBM Active**<br>**Control, N = 48<sup>a</sup>** | **A-ApBM**<br>**Intervention, N = 48<sup>a</sup>** | **p-**<br>**value** |
| --- | --- | --- | --- | --- |
| **Sex** |  |  |  | 0.5<sup>b</sup> |
| Female | 10 / 40 (25%) | 12 / 48 (25%) | 8 / 48 (17%) |  |
| Male | 30 / 40 (75%) | 36 / 48 (75%) | 40 / 48 (83%) |  |
| **Age** | 31.71 (8.34) | 33.78 (5.32) | 34.51 (6.20) | 0.14<sup>c</sup> |
| **Dominant Hand** |  |  |  | 0.7<sup>b</sup> |
| Left | 4 / 40 (10%) | 7 / 48 (15%) | 8 / 48 (17%) |  |
| Right | 36 / 40 (90%) | 41 / 48 (85%) | 40 / 48 (83%) |  |
| **Marital Status** |  |  |  | 0.7<sup>b</sup> |
| Divorced | 10 / 40 (25%) | 14 / 48 (29%) | 14 / 48 (29%) |  |
| Married | 15 / 40 (38%) | 19 / 48 (40%) | 13 / 48 (27%) |  |
| Unmarried | 15 / 40 (38%) | 15 / 48 (31%) | 21 / 48 (44%) |  |
| **Education** |  |  |  | 0.2<sup>b</sup> |
| College or<br>Above | 3 / 40 (7.5%) | 0 / 48 (0%) | 4 / 48 (8.3%) |  |
| Elementary<br>School | 3 / 40 (7.5%) | 3 / 48 (6.3%) | 3 / 48 (6.3%) |  |
| Junior High<br>School | 22 / 40 (55%) | 26 / 48 (54%) | 32 / 48 (67%) |  |
| Senior High<br>School | 12 / 40 (30%) | 19 / 48 (40%) | 9 / 48 (19%) |  |
| **Smoker** | 32 / 40 (80%) | 43 / 48 (90%) | 43 / 48 (90%) | 0.3<sup>b</sup> |
| **Drinker** | 24 / 40 (60%) | 27 / 48 (56%) | 37 / 48 (77%) | 0.078<sub>b</sub> |
| **Meth Use** | 2.80 (2.40) | 2.55 (2.25) | 3.17 (2.87) | 0.6<sup>c</sup> |

| Characteristics | No-intervention Control, N = 40<sup>a</sup> | S-ApBM Active Control, N = 48<sup>a</sup> | A-ApBM Intervention, N = 48<sup>a</sup> | p-value |
| --- | --- | --- | --- | --- |
| **History (yrs)** |  |  |  |  |
| **Heroin History** | 1 / 40 (2.5%) | 1 / 48 (2.1%) | 2 / 48 (4.2%) | 0.8<sup>b</sup> |
| **Ketamine History** | 0 / 40 (0%) | 1 / 48 (2.1%) | 0 / 48 (0%) | 0.4<sup>b</sup> |
| **Craving (baseline)** | 7.66 (11.79) | 6.33 (9.28) | 8.65 (13.44) | 0.8<sup>c</sup> |

<sup>a</sup>n / N (%); Mean (SD)
<sup>b</sup>Pearson's Chi-squared test
<sup>c</sup>Kruskal-Wallis rank sum test

Figure 1 displays the CONSORT flow diagram illustrating the participants’ progression throughout the study.

**Figure 1:**
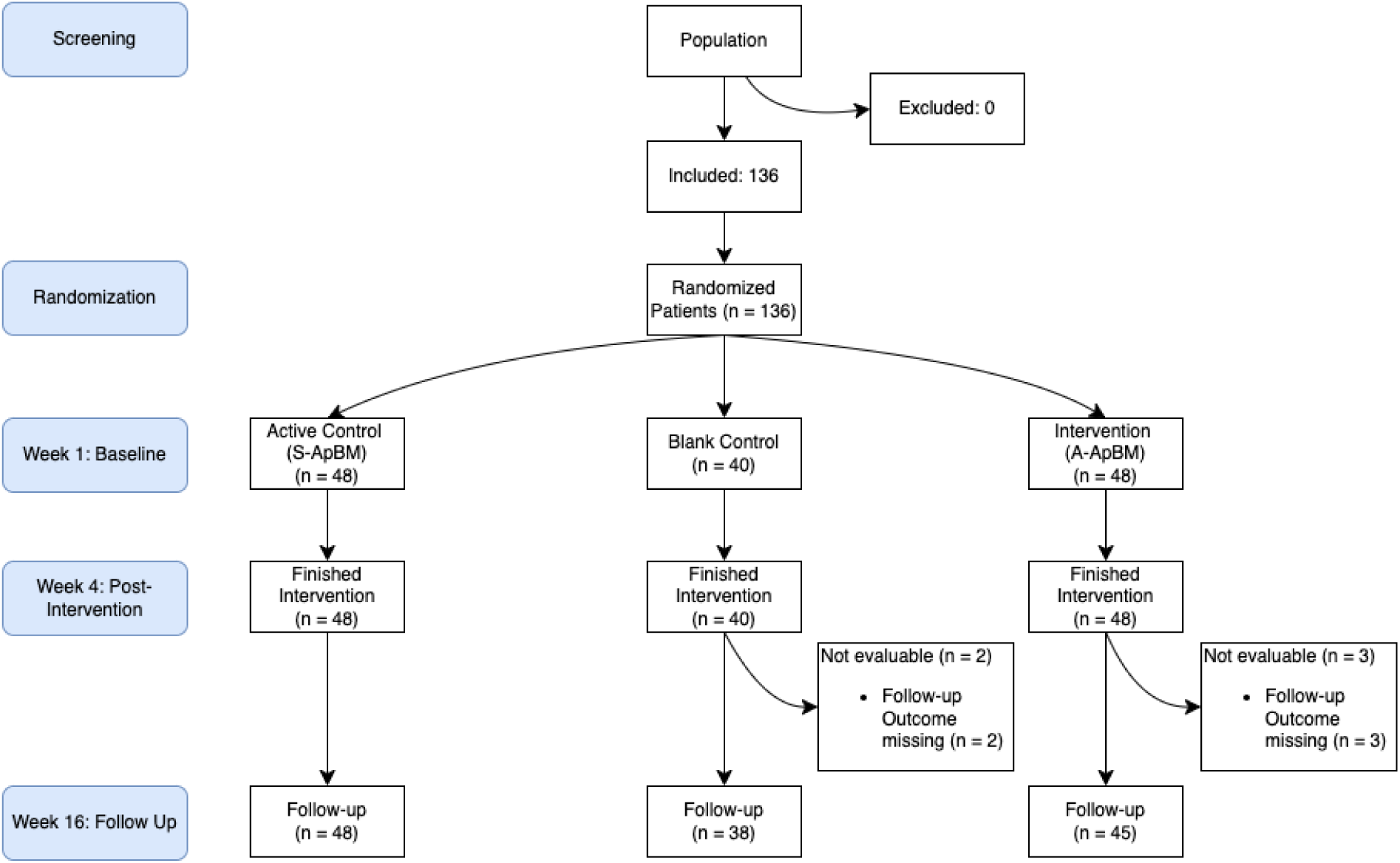
CONSORT Diagram

**Figure 2:**
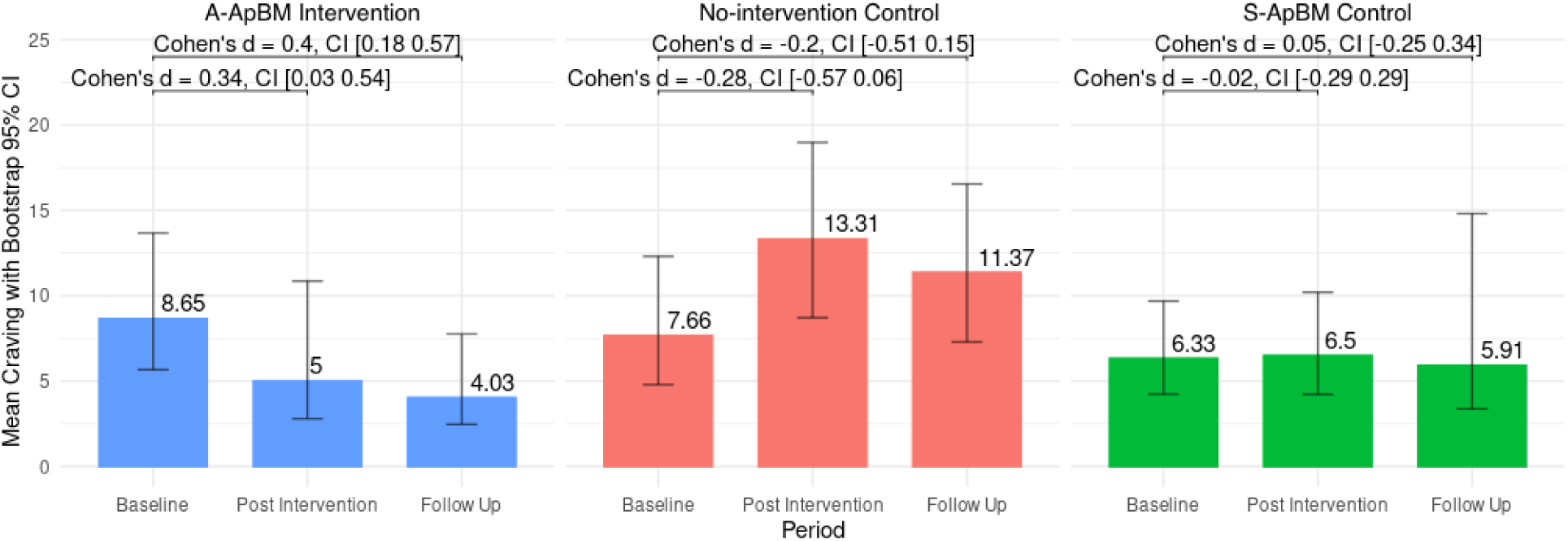
Change in Cue-Induced Craving

### Training Time

We calculated the daily training time for each user, which is calculated by dividing the total duration of time each user spent in the training by the number of days that each user was engaged in the training. The A-ApBM group had an average daily training time of 5.32 minutes (SD = 1.43 minutes), and the S-ApBM group had an average daily training time of 5.09 minutes (SD = 1.53 minutes).

### Change in Cue-induced Craving

There was a statistically significant interaction between group and time in explaining the craving score, F(4,266) = 3.02 (95% CI = [1.18,11.20]), p= 0.018, 1^2^ = 0.043 (95% CI = [0.016,0.141]). Bonferroni-corrected pairwise comparison showed that the mean craving score was significantly different at Post Intervention between No-intervention Control and A- ApBM (p = 0.01) and between No-intervention Control and S-ApBM (p = 0.05). At Follow Up the mean craving score was significantly different between No-intervention Control and A- ApBM (p = 0.03). Bootstrapped pairwise t-tests revealed that only the A-ApBM group showed differences between Baseline and Post Intervention (Cohen’s d = 0.34, p< 0.01, 95% CI = [0.03,0.54]) and between Baseline and Follow Up (Cohen’s d = 0.40, p = 0.01, 95% CI = [0.18,0.57]).

### Relapse

Throughout the study duration, including at the 4-week and 16-week follow-up points, there were no instances of self-reported relapse among participants across all study groups. This observation was further corroborated by objective measures: urine drug tests conducted at weeks 4 and 16 yielded uniformly negative results for all participants, irrespective of their group assignment.

### Relationship between Performance and Difficulty Indices

Table 2 tabulates the estimates of the fixed effects of the linear mixed-effects model. The explanatory power is substantial (conditional R^2^ = 0.82). The 95% Confidence Intervals and p-values were computed using a Wald t-distribution approximation. The effect of ITR was significant and positive (Beta = 2.18, p = 0.027), indicating that as the ITR increased, participants’ performance improved. The quadratic term for ITR was negative and highly significant (Beta = -2.47, p < 0.001), suggesting an inverted U-shaped relationship between ITR and performance. This means that performance improved with increasing difficulty up to a certain point, after which further increases in difficulty resulted in diminished performance, supporting the theory that moderate difficulty is most effective for learning. Time (t) also had a significant and positive effect (Beta = 2.95, p < 0.001), indicating that performance improved over time as participants became more familiar with the task. The non-significant quadratic term for time (p = 0.7) suggests that while participants’ performance improved over time, the learning curve did not exhibit a significant acceleration or deceleration during the intervention. Lastly, the standard deviation of the ITR from recent sessions (Past3ITRSD) was negatively associated with performance, although this effect was not statistically significant (p = 0.10). This indicates that greater variability in difficulty across sessions tended to reduce performance, though this finding warrants further investigation.

**Table 2:** Mixed Effect Regression Results.

| Characteristic | Beta<sup>1</sup> | 95% CI<sup>2</sup> | p-value |
| --- | --- | --- | --- |
| ITR |  |  |  |
| ITR | 2.18* | 0.25, 4.1 | 0.027 |
| ITR<sup>2</sup> | -2.47*** | -3.6, -1.3 | <0.001 |
| $t$ | | | |
| $t$ | 2.95*** | 2.2, 3.8 | <0.001 |
| $t^2$ | 0.13 | -0.58, 0.84 | 0.7 |
| Past3ITRSD | -6.30 | -14, 1.3 | 0.10 |

<sup>1</sup> \*\*\*: $p < 0.001$ , \*\*: $p < 0.01$ , \*: $p < 0.05$ ; <sup>2</sup> CI = Confidence Interval

## Discussion

The findings of this pilot randomized controlled trial are twofold. First, there was a significant reduction in craving scores in the A-ApBM group and a lack of significant change in the control groups. Second, a significant relationship between performance and difficulty indices within the A-ApBM intervention indicates the merit of the proposed adaptive algorithm. These findings underscore the potential effectiveness of an adaptive approach bias modification (A- ApBM) intervention in reducing cue-induced cravings among individuals with a history of methamphetamine use.

One of the primary outcomes observed was a significant reduction in craving scores in the A-ApBM group, both immediately after the intervention and at the 16-week follow-up. This suggests that the A-ApBM intervention may have a lasting impact, which could be particularly beneficial for interventions aimed at behavior change or addiction treatment. The observed reduction in cravings is likely attributable to the adaptive and personalized nature of the A- ApBM intervention, which dynamically adjusts the difficulty level based on the user’s performance. This approach appears to maintain engagement and challenge, potentially enhancing the overall effectiveness of cognitive bias modification (CBM) programs.

In contrast, the control groups, including the static approach bias modification (S-ApBM) group and the no-intervention control group, did not exhibit significant changes in craving scores over time. The use of an active control like S-ApBM is crucial in isolating the effect of specific intervention components. The lack of significant findings in the S-ApBM group suggests that the specific elements of the A-ApBM intervention, rather than mere participant engagement, may be driving the observed effects. This lack of significant change in the control groups highlights the importance of the specific elements incorporated into the A-ApBM intervention. The static nature of the S-ApBM, which did not adapt to individual performance, was insufficient to produce a meaningful reduction in cravings, emphasizing that engagement alone, without the dynamic adjustment of training difficulty, may not be enough to achieve the desired therapeutic outcomes.

Moreover, the study revealed a significant relationship between performance and difficulty indices within the A-ApBM intervention. The adaptive algorithm used in this study was designed to maintain an optimal level of difficulty by continuously adjusting based on real-time performance data. This relationship between performance and difficulty underscores the importance of personalized interventions in maintaining user engagement and enhancing training efficacy. By leveraging this relationship, the adaptive algorithm helps to ensure that the training remains challenging yet achievable, thereby maximizing its therapeutic impact.

These findings have broader implications for the development of future interventions for substance use disorders and other psychiatric conditions. The success of the A-ApBM intervention in this study suggests that incorporating gamification and adaptive elements into CBM programs may be a promising strategy for improving outcomes in various therapeutic contexts. The effect size, measured by Cohen’s d, suggests a small to moderate effect, which could imply that patients might continue to benefit even after the intervention has ended. This could reduce the need for continuous treatment and potentially lower the costs and resource use in clinical settings. Future research could explore the application of similar adaptive algorithms in other types of addiction treatments or in interventions targeting different psychiatric disorders.

Previous study [25] have found that monotony, repetition, lack of novelty, and lack of complexity cause boredom. Boredom can negatively impact engagement, attention, and interest, which can ultimately reduce the effectiveness of CBM programs. The mixed-effect linear regression indicates that the posited relationship between difficulty and performance was evident.

The inverted U-shape effect of the Individualized Task Difficulty on performance and the negative correlation with the standard deviation of past difficulty indices suggest that the algorithm makes the ApBM training more engaging, effective, and personalized for individuals with substance use disorder. This adaptive approach has the potential to improve treatment outcomes and contribute to the field of clinical psychology and addiction treatment.

## Limitations

We acknowledge the following limitations of the study. First, the sample size was relatively small and homogenous in race and ethnicity, which may limit the generalizability. The originally planned sample size of 300 participants across six groups was revised to 150 participants across three groups due to logistical constraints and feasibility considerations.

Specifically, the limited number of eligible individuals in the rosters of the 12 participating community rehabilitation centers necessitated this adjustment. While we successfully enrolled 136 participants, nearly reaching our revised target, the recruitment phase was concluded when the available pool of eligible participants was exhausted. This reduction in sample size may limit the generalizability of our findings. Furthermore, the decision to exclude additional algorithmic variants (U-shape, N-shape, and inverted N-shape difficulty curves) reduced the scope of the study. Future research should aim to replicate these findings with larger sample sizes and include evaluations of the excluded algorithmic variants to validate and expand upon our results. Second, the duration of the intervention program was relatively short at 4-week intervention and 16-week follow up. Third, the relapse outcome might have been influenced by participant behavior due to their awareness of the study. Additionally, the loss of five participants from the control and A- ApBM groups at week 16, who did not complete the follow-up assessments, raises unconfirmed concerns about possible relapse. This attrition and the potential for behavior modification among participants highlight the need for longer-term studies with strategies to ensure sustained engagement for a more accurate assessment of relapse prevention in methamphetamine use. In considering the relapse rates observed in our study, it is important to contextualize these findings within the framework of participant behavior and study design. The participants were aware that the study duration was 4 weeks, which might have influenced their commitment to abstain from methamphetamine use during this period. This awareness could have motivated participants to consciously refrain from relapsing, in an effort to demonstrate their engagement and adherence to the study protocols. Additionally, it is noteworthy that in both the no intervention control group and the A-ApBM group, there were instances of participant attrition at the 16-week mark— two and three participants, respectively, were lost to follow-up and did not complete the craving assessment or the urine tests. While the reasons for this attrition are unknown, one cannot rule out the possibility of relapse among these individuals. However, this remains speculative in the absence of concrete evidence. The loss of these participants from the follow-up assessment phase underlines the complexities involved in conducting long-term studies in populations with substance use disorders and highlights the potential challenges in maintaining consistent participant engagement over extended periods.

## Conclusion

In conclusion, this study highlights the promise of an adaptive and gamified approach to cognitive bias modification in mitigating cue-induced cravings among individuals with a history of methamphetamine use. The dynamic adjustment of training difficulty based on individual performance is crucial in maximizing intervention effectiveness. These findings emphasize the need for engagement and personalization in CBM programs, suggesting that adaptive strategies could significantly enhance treatment outcomes for substance use disorders. Future research should continue to explore these adaptive elements to further validate their benefits in diverse therapeutic contexts.

## Data Availability

All data produced in the present study are not available

## Acknowledgments

We would like to thank the participating social workers and administrative staffs at the participating communities.

## Data Availability

A example data and code for replicating the algorithm is available at https://github.com/liuxiang91/A-ApBM/.

## Author Contributions

The authors made the following contributions. Liqun Zhang, BS: Conceptualization, Writing - Original Draft Preparation, Investigation, Data curation, Software; Yanru Liu, MS: Writing - Original Draft Preparation, Writing - Review & Editing; Xiang Liu, PhD: Conceptualization, Formal analysis, Validation, Supervision; Yuanhui Li, MS: Conceptualization, Validation, Software; Tianjiao Zhang, MS: Data curation, Software; Dai Li, BS: Conceptualization, Project administration; Danlin Shen, MD and Jianping Jiao, BS and Wei Hao, MD: Supervision, Writing - Review & Editing.

## Abbreviations

CBM: CognitiveBias Modification
ApBM: Approach Bias Modification
A-ApBM: Adaptive Approach Bias Modification
S-ApBM: Static Approach Bias Modification
MUD: Methamphetamine Use Disorder
ITR: Intended Training Ratio

